# Impact of preterm premature rupture of membranes on composite adverse perinatal outcomes and associated predictors among pregnant women in Tigray, northern Ethiopia: a prospective cohort study

**DOI:** 10.64898/2026.02.22.26346847

**Authors:** Girmay Teklay Welesamuel, Aregawi Araya, Gebreab Nega, Belay Alem, Teklay Guesh, Hareg Mekonen, Fiseha Abadi, Hagos Gebreluel, Negasi Asress, Teklehaimanot Gereziher Haile, Tsega Teshale Alemayoh

**Affiliations:** Department of Pediatrics and Child health nursing, school of nursing, Aksum university, Ethiopia; Department of Maternity and Neonatal Nursing, school of nursing, Aksum university, Ethiopia; Department of Pediatrics and Child health, school of Medicine, Aksum university, Ethiopia; Department of Gynecology and Obstetrics, school of Medicine, Aksum university, Ethiopia; Department of Pediatrics ward, comprehensive specialized hospital, Aksum university, Ethiopia; Department of Biostatics and Epidemiology, school of public health, Aksum university, Ethiopia; Department Medical Laboratory, Aksum university, Ethiopia

**Keywords:** Preterm Premature Rupture of Membrane, composite adverse perinatal outcomes, latency period, predictors, Tigray, Ethiopia, prospective cohort

## Abstract

**Background:** Preterm premature rupture of membranes (PPROM) is a leading contributor of adverse perinatal outcomes, particularly in low-resource and conflict-affected settings. Despite its clinical importance, prospective evidence on its impact on composite adverse perinatal outcomes in northern Ethiopia remains limited. This study examined the impact of Preterm premature rupture of membranes on composite adverse perinatal outcomes and identified associated predictors among pregnant women in public hospitals of Tigray, Northern Ethiopia.

**Methods:** A hospital-based prospective cohort study was conducted among 578 singleton pregnancies (288 with Preterm premature rupture of membranes and 290 without it at ≥28 weeks of gestation. Participants were followed from admission to delivery and to the early neonatal period. The primary outcome was a Composite adverse perinatal outcome, and the main exposure variable was Preterm premature rupture of membranes (PPROM). Modified Poisson regression with robust variance estimation was used to estimate adjusted relative risks (ARRs) with 95% confidence intervals (CIs) and a significant level was declared at p<0.05.

**Results:** Overall, 33.4% of neonates experienced at least one composite adverse perinatal outcome. The incidence was substantially higher among the PPROM group compared with the non-PPROM group (59.4% vs. 7.6%). After adjustment, PPROM was strongly associated with composite adverse perinatal outcomes (ARR = 7.22, 95% CI: 4.73-11.03). Independent predictors included previous pregnancy-related infection (ARR = 1.54; 95% CI: 1.08–2.22), absence of iron-folate supplementation during pregnancy (ARR=1.63; 95% CI: 1.153-2.29), pelvic pain (ARR = 2.09; 95% CI: 1.05–4.15), and latency period of 1–3 days (ARR = 1.41; 95% CI: 1.10–1.81) compared to <24 hours. Induced labor was protective (ARR=0.58; 95% CI: 0 .422-0.800).

**Conclusion:** PPROM markedly increases the risk of composite adverse perinatal outcomes in this post-conflict, resource-constrained setting. The first 72 hours following membrane rupture represent a particularly vulnerable period. Strengthening antenatal care, nutritional supplementation, infection prevention, and timely obstetric intervention could reduce preventable neonatal morbidity and mortality in similar contexts.

## Introduction

Preterm premature rupture of membranes (PPROM) is a major obstetric complication characterized by rupture of the fetal membranes before commencement of labor and prior to 37 completed weeks of gestation(1). In Ethiopia, PPROM is defined as membrane rupture occurring after fetal viability (≥28 weeks of gestation) but before 37 weeks of gestation (2).

Globally, PPROM is a significant contributor to preterm birth and adverse perinatal outcomes (3). It affects approximately 3% of all pregnancies and accounts for one-third of preterm deliveries(4, 5). Overall, preterm birth affects approximately 11% of live births and remains a leading Cause of neonatal morbidity and death, notably in sub-Saharan Africa(6–8).

Neonates delivered following PPROM are especially vulnerable due to the combined effects of prematurity, prolonged intrauterine exposure after membrane rupture, ascending infection, and inflammatory processes (3, 9). These neonates are at increased risk of respiratory distress syndrome (RDS), sepsis, low Apgar scores, intraventricular hemorrhage, necrotizing enterocolitis, NICU admission, and early mortality, often occurring concurrently (3, 10, 11). Therefore, Composite adverse outcomes better capture this cumulative burden(12, 13).

The impact of PPROM is pronounced in low- and middle-income countries, particularly in sub-Saharan Africa (14, 15), where healthcare infrastructure and neonatal care are often limited (16–19). In Ethiopia, the national prevalence of PPROM is estimated at 9.2% (20, 21) while higher rates ranging from 12.5% to 13.7% have been reported in the Tigray region (21–23).These outcomes are further aggravated by prematurity (3, 24), lengthy latency periods(25–27), and maternal infections (28).

In the Tigray region, the persistent challenges are due to the shortage of providers, delay referral, and inadequate neonatal care capacity (29–38), which have been compounded by the 2020-2022 conflict, during which more than 70% of health facilities were damaged during (39). The disruption of health services supply chains and delayed referral networks has further intensified the risks associated with PPROM in the post-conflict setting (15, 40–42).

Although efforts have been made to improve maternal and neonatal care, adverse neonatal outcomes related to PPROM remain high in Ethiopia. Most existing studies are retrospective and focused on isolated outcomes (15, 22, 43, 44). Prospective evidence on composite adverse outcomes in northern Ethiopia is scarce, restricting the development of targeted and context-specific interventions.

Therefore, this study aimed to assess the association between PPROM and composite adverse perinatal outcomes among neonates in Tigray’s public hospitals using a prospective cohort design, comparing PPROM and non-PPROM groups to inform clinical practice, healthcare strengthening, and policy in resource-limited, post-conflict settings.

## Methods

### Study Design and Setting

A hospital-based prospective cohort study was conducted to evaluate the impact of preterm premature rupture of membranes (PPROM) on composite adverse perinatal outcomes. The study was carried out in selected public hospitals of the Tigray region of northern Ethiopia. These hospitals serve as major referral centers for maternal and neonatal care, providing antenatal, intrapartum, and neonatal intensive care services for both urban and rural populations.

### Study Population

The source population comprised all pregnant women who delivered in the selected public hospitals during the study period. From this group, two cohorts were prospectively established. The study population included pregnant women from 28 weeks of gestation who came to the hospital. The exposed cohort (PPROM group) consisted of women with singleton pregnancies who had a confirmed diagnosis of preterm premature rupture of membranes before the onset of labor. The unexposed (comparison) cohort consisted of women with singleton pregnancies who delivered without any diagnosis of PPROM, regardless of their gestational age at delivery

### Eligibility Criteria

Women were eligible to participate if they carried a singleton pregnancy. For the PPROM group, confirmation of PPROM before labor was required. For the comparison group, delivery had to occur without PPROM. We excluded multiple pregnancies (twins or higher), fetuses with major congenital anomalies known to be incompatible with life, and any mothers who declined to give consent or who were lost to follow-up before the baseline assessment could be completed.

### Sample Size and Sampling Technique

The sample size was calculated using assumptions of 95% confidence level, 90% power, and a 1:1 ratio of exposed to non-exposed groups.

Gestational age at delivery was used as a predictor with an expected outcome proportion of the unexposed group 55% and an odds ratio of 1.8 were used from a previous study(15). Considering a 10% non-response rate, a total sample size of 605 mother–neonate pairs was included (303 in the PPROM group and 302 in the non-PPROM group).

A consecutive sampling technique was used to enroll eligible women with PPROM as they presented to the hospitals. For each enrolled PPROM case, we immediately selected one woman delivering without PPROM in the same hospital during the same period to serve as her matched comparison participant. This approach ensured that both groups were recruited from the same facilities under similar conditions. Participant recruitment for this prospective cohort study was conducted from 2 November 2024 and ends 10 October 2025. All participants were followed prospectively from the time of hospital admission, through delivery, and into the early neonatal period.

### Data Collection Procedures

Data were collected using three main sources. A structured interviewer-administered questionnaire to gather maternal socio-demographic and obstetric history, direct review of medical records for intrapartum and delivery details (2/11/2024 to 10/10/2025), and a standardized neonatal follow-up form completed by trained midwives and neonatal nurses to document birth and early neonatal outcomes.

The data collection tools were originally prepared in English, translated into Tigrigna, and back-translated to English to ensure accuracy and cultural appropriateness. All data were captured electronically using the Kobo Toolbox platform, which allowed real-time validation checks and reduced errors during entry.

### Variables and measurements

#### Outcome variable

The primary outcome was composite adverse birth outcome, defined as the occurrence of at least one of the following conditions identified at birth or early neonatal period:

❖ Low Apgar score at 5 minutes (<7)
❖ Respiratory distress at birth
❖ Need for immediate neonatal resuscitation
❖ Admission to the neonatal intensive care unit (NICU) immediately after birth
❖ Stillbirth or early neonatal death occurring at delivery

### Exposure variables

The main exposure variable was the presence or absence of PPROM. We also collected a wide range of potential confounding and mediating variables, including maternal age, parity, educational level, antenatal care attendance, gestational age at delivery, mode of delivery, history of maternal infections, use of iron-folate supplementation, duration of the latency period (time from membrane rupture to delivery), presence of pelvic pain, and previous obstetric complications.

### Data Quality Assurance

Several steps were taken to ensure high data quality. All data collectors and field supervisors received intensive standardized training before the study began. The data collection tools were pretested in a non-study hospital and revised based on feedback before the actual data collection time. Supervisors reviewed completed forms daily for completeness and logical consistency. Regular on-site supervision visits and real-time data monitoring through the electronic platform helped maintain accuracy throughout the study period.

### Ethical Considerations

Ethical approval was obtained from the Health Research Review committee (HRERC) of Aksum University College of Health Sciences and specialized hospital with approval number of 075/2024 on November 23, 2024. Recruitment of data collector was started directly after getting support letter from Tigray Regional Health bureau on 02 December, 2024. An approval letter was also obtained from participating hospitals. Written informed consent was obtained from all participants. Confidentiality was maintained throughout the study, and participation was voluntary.

### Data Analysis

Data were collected electronically using Kobo Toolbox, exported to STATA version 16 for cleaning and analysis. Descriptive statistics were used to summarize maternal, obstetric, infection-related, delivery, and neonatal characteristics. Categorical variables were described using frequencies and percentages, while continuous variables were summarized using means and standard deviations. Comparisons between mothers with preterm premature rupture of membranes (PPROM) and those without PPROM were performed using Chi-square tests for categorical variables and independent t-tests for continuous variables.

To estimate the effect of PPROM on composite adverse perinatal outcomes, modified Poisson regression with robust variance estimation was employed. This approach was selected to directly estimate relative risks (RRs) due to the convergence issue of the log binomial model. Robust standard errors were applied to account for potential overdispersion and to provide valid confidence intervals. In the bivariable analysis, crude relative risks (RRs) with 95% confidence intervals were calculated for each independent variable. Variables with a p-value <0.25 in the bivariable analysis, along with variables identified a priori based on clinical and epidemiological relevance, were included in the multivariable modified Poisson regression model. Model adequacy was assessed using goodness-of-fit measures and by examining variance inflation factors to evaluate multicollinearity among covariates. Results from the multivariable analysis were reported as adjusted relative risks (ARRs) with 95% confidence intervals (CIs) and statistical significance was declared at a p-value <0.05.

## Result

### Participant Flow and Follow-up

A total of 605 pregnant women were enrolled in the cohort, comprising 303 women with PPROM and 302 without PPROM. After excluding twenty-seven participants with incomplete or missing key information, 578 mother–neonate pairs were included in the final analysis (288 in the PPROM group and 290 in the non-PPROM group), yielding a response rate of 95.5%. All enrolled participants were followed from admission through the immediate postnatal period to assess perinatal outcomes (**Fig 1**).

**Fig1.**
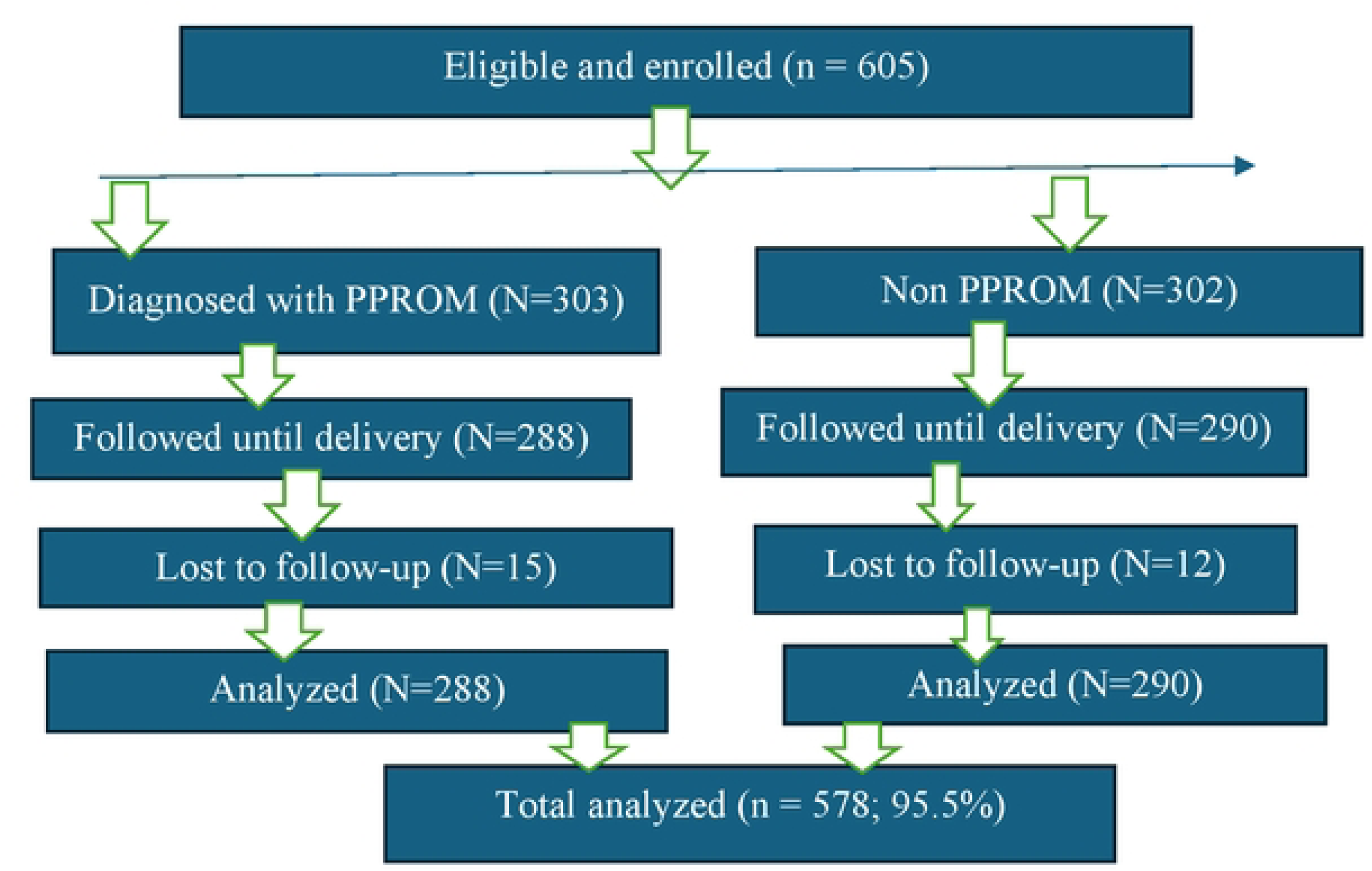
Flow diagram of participant recruitment, follow-up, and analysis for the study on the impact of preterm premature rupture of membranes on composite adverse perinatal outcomes in Tigray, Northern Ethiopia

### Socio-demographic characteristics

The majority of participants were within the prime reproductive age group, with a mean maternal age of 27.8 ± 5.8 years (17–46 years). Most mothers (98.96%) were married. A majority lived in urban areas (61.2%), although rural mothers experienced a higher proportion of composite adverse perinatal outcomes (35.7%) compared with urban mothers (31.9%). Lower maternal education was linked to poorer outcomes. About 80% of neonates with adverse perinatal outcomes were born to mothers who had only primary education or no formal schooling. Housewives formed the largest occupational group (58.13%) and accounted for 65% of adverse outcomes, while employed women and daily laborers had lower rates.

### Previous obstetrics and medical history

Looking at previous obstetric history, 9.34% of mothers reported a prior preterm birth, and 41.2% of these experienced adverse perinatal outcomes in the current pregnancy. Among the 30 women with a history of stillbirth, 53.3% had composite adverse outcomes in the index pregnancy. A previous history of PPROM (reported by 36 mothers) showed one of the strongest associations, with 72.2% of these women having adverse perinatal outcomes.

Previous pregnancy complications of any type were documented in 42 mothers, of whom 47.6% (20/42) had adverse perinatal outcomes, compared to 32.3% (173/536) among those without complications. A history of any pregnancy-related infection, although rare (only 10 mothers), was associated with the highest proportion of adverse outcomes (80%). A history of miscarriage or spontaneous/induced abortion was reported by 84 mothers, with 38.1% (32/84) experiencing composite adverse outcomes. Other factors such as previous cesarean section (especially emergency), admission to neonatal intensive care, low birth weight in a prior child also appeared more frequently among women who experienced poor outcomes in the current pregnancy.

### Obstetric and Clinical Characteristics of the Current Pregnancy

The overall incidence of composite adverse perinatal outcomes was substantially higher among neonates born to mothers with PPROM compared to those without PPROM (59.4% vs. 7.6%, respectively). From the total 160 mothers whose MUAC less than 23 cm, 104(65%) had adverse perinatal outcome. Self-reported fluid leakage before the onset of labor was associated with 36.0% adverse outcomes versus 11.3% in those without leakage. Among PPROM cases, 97.9% (282/288) received immediate management, most commonly antibiotics (48.44%) followed by corticosteroids (41.87%).

Earlier gestational age at membrane rupture was strongly associated with adverse outcomes: 77.9% poor outcomes at 28–33+6 weeks, 52.5% at 34–36+6 weeks, and 6.9% at ≥37 weeks. Delayed presentation for care-seeking after membrane rupture (>6 hours) was linked to a higher proportion of poor outcomes (40.2%). With respect to satisfaction with healthcare providers, adverse outcomes were reported in 51 (35.4%) mothers reporting good satisfaction.

Other factors with notable proportions of composite adverse perinatal outcomes included primiparity (40.3%), birth interval <23cm (39.8%), non-cephalic presentation (47.3%), history of urinary tract infection (35.5%), antepartum hemorrhage (33.8%), pre-eclampsia/eclampsia (32.6%), anemia (HGB <11 g/dl; 33.5%), vaginal bleeding (33.1%), and carrying heavy objects during pregnancy (35%).

### Maternal Health-seeking behavior

Women who reported feeling uncomfortable discussing their health issues had a markedly higher proportion of composite adverse perinatal outcomes (66.7%) compared to those who felt comfortable (33.0%). Similarly, those dissatisfied with the information provided during care showed higher rates of poor outcomes (66.7% vs. 33.0%).

Skipping antenatal care appointments was associated with a slightly higher proportion of adverse outcomes, 35.8% of cases, compared to 31.8% among those who attended all appointments.

Among women who skipped appointments, fear of diagnosis was linked to the highest rate of poor outcomes (57.1%), followed by financial issues (36.3%), distance to the facility (35.8%), and lack of transportation (33.5%).

Choice of healthcare service based on recommendations from family or friends was linked to a higher proportion of adverse outcomes (45.0%), while proximity to home showed the lowest (30.6%). Lack of family support for seeking healthcare and discomfort in discussing health, though based on small numbers 2(25.0%), also suggested poorer outcomes.

### Infection-Related Clinical Factors

Clinical indicators of infection were among the strongest predictors of composite adverse perinatal outcomes. Among 162(28%) mothers with PV over examination (>=5times), 56(34.6%) of them were exposed to adverse perinatal outcome. Mothers with foul-smelling vaginal discharge were associated with 88.9% adverse outcomes compared to 31.6% in its absence. Pelvic pain during the current pregnancy showed 50.0% adverse outcomes versus 33.0% without pain. About 494(85.5%) who received contact information for emergency, 169(34.2%) of them were exposed to ana adverse perinatal outcome.

### Delivery characteristics and immediate neonatal outcomes

Delivery and immediate neonatal variables showed clear gradients with composite adverse perinatal outcomes. The mean of duration of labor in hours was 10.6 (SD ±9.7), while the mean birth weight was 2877.5 g (SD ±607.8) ranging from 1200 g to 4500 g. Latency period duration strongly influenced outcome. Adverse perinatal outcome occurred in 49(22.1%) with shortest latency (<24 hours), increased to 56(50%) among those with latency exceeding seven days. About 36(20.9%) of neonates developed adverse perinatal outcome after induced labor onset compared to spontaneous onset 157(38.7%).

Extremely and moderate preterm neonates 28–33+6 weeks experienced 32(84.2%) adverse outcomes, while those delivered at 34–36+6 weeks had a 125(75.3%) rate. Term neonate at ≥37 weeks had 36(9.6%) adverse outcomes. Small-for-gestational-age neonates also had 56(68.3%) adverse outcomes.

Immediate postnatal interventions were highly indicative of adverse outcome. Neonatal requiring resuscitation had 87(80.6%) rates of adverse outcome. Similarly, 31(70.5%) neonates with delivery complications experience with adverse outcome, with the most common complications being placental abruption, previa, and meconium aspiration syndrome.

### Perinatal outcome

Overall, one third of the neonates (33.4 %) experienced composite adverse perinatal outcomes. At delivery, the most immediate adverse conditions were low birth weight accounting for 87 (45%), followed by prematurity in 80(41.5%) neonates. Stillbirth occurred in 8(4.2%) neonates, representing a small but clinically significant proportion of adverse outcome (**Fig 2**).

**Fig 2:**
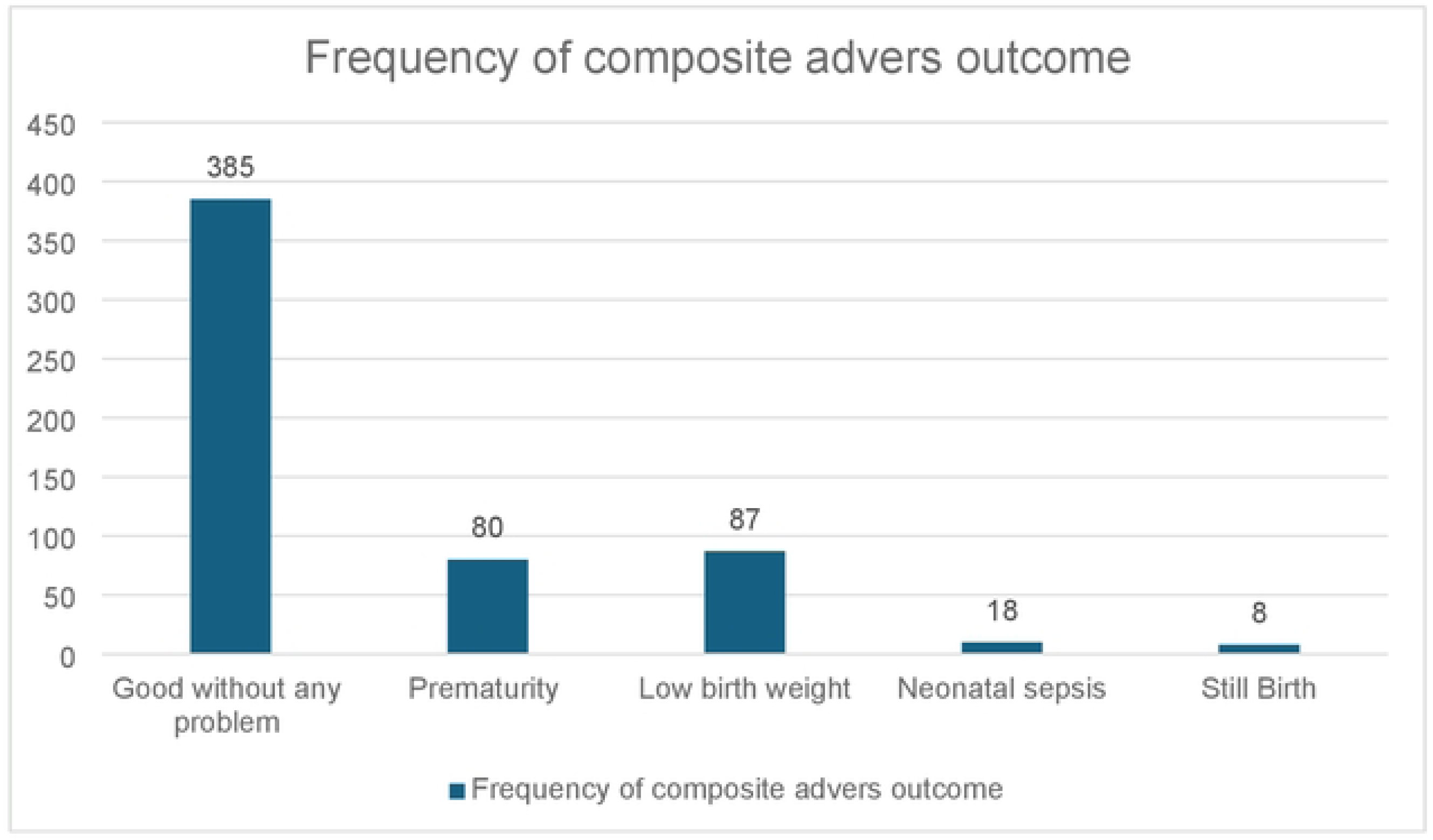
Composite Adverse perinatal outcome among study participants in Tigray region, northern Ethiopia

During the early neonatal period, indicators of compromised neonatal status were strongly associated with adverse perinatal outcome. A low APGAR score (4–7) at the first minute was substantially more frequent among neonates with adverse outcomes. Of the 98 neonates with a 1-minute APGAR score of 4–7, 80(81.6%) developed adverse perinatal outcomes. The association was even more pronounced at five minutes. Among neonates with a low APGAR score (4–7), 48(96%) developed adverse outcomes, underscoring the prognostic significance of delayed physiological recovery after birth.

Similarly, the need for immediate clinical intervention was highly predictive of poor outcomes. Among neonates who required respiratory support during delivery, 80 (90.9%) of them developed adverse outcomes. A total of 122 neonates were admitted to the NICU. Among them, 110(90.2%) had adverse perinatal outcomes. The leading indications for NICU admission were Low birth weight, Respiratory distress syndrome, Feeding difficulties and related neonatal complications.

### Effect of PPROM on Composite Adverse Perinatal Outcomes

Among the whole participant 288(49.83%) were PPROM mothers. Among them 171(59.4%) had composite adverse Perinatal outcome. After adjustment for potential confounders, PPROM remained the strongest independent predictor of composite adverse perinatal outcomes. Neonates born to mothers with PPROM had more than seven times higher risk of experiencing at least one adverse outcome compared to those without PPROM (ARR = 7.22; 95% CI: 4.73–11.03; p<0.001).

## Discussion

This prospective cohort study assessed the effect of preterm premature rupture of membranes (PPROM) on composite adverse perinatal outcomes in public hospitals of the Tigray region, northern Ethiopia. Preterm premature rupture of membranes (PPROM) was the strongest independent predictor of composite adverse perinatal outcomes in this study. After adjustment for confounding variables, PPROM increased the risk of adverse neonatal outcomes more than sevenfold (ARR = 7.22; 95% CI: 4.73–11.03). This magnitude of association indicates a substantial and independent contribution of PPROM to neonatal morbidity in this population.

The association is biologically plausible, as membrane rupture compromises the intrauterine protective barrier (3), facilitating ascending infection, chorioamnionitis, and fetal inflammatory response(10), which contribute to neonatal sepsis, respiratory distress syndrome (RDS) (45), bronchopulmonary dysplasia, and early neonatal mortality(15, 43, 46). Similar findings have been reported in Ethiopian and other African cohort studies (3, 47). The pronounced disparity observed in this study underscores the profound impact of membrane rupture on neonatal health, particularly within resource-limited and post-conflict settings (15, 48).

This elevated risk likely reflects the combined effects of prematurity, intrauterine inflammation, delayed care-seeking (with 40.2% poor outcomes among those presenting >6 hours after rupture), and limited neonatal intensive care capacity within the post-conflict context of Tigray (49, 50). Consistent evidence from other low- and middle-income countries suggests that health system capacity plays a critical role in modifying PPROM-related outcomes (10, 15, 42). In contrast, settings with timely obstetric interventions, well-established referral systems, and adequately equipped neonatal intensive care units report lower complication rates, suggesting that health system capacity plays a critical modifying role (51).

Regarding the latency period (time from rupture of membranes to delivery), our findings demonstrate a critical non-linear relationship (25, 27, 52). Compared with delivery within 24 hours, a latency of 1–3 days was associated with a 41% increased risk of composite adverse perinatal outcomes (ARR = 1.41; 95% CI: 1.10–1.81), whereas a latency period of 4–7 days demonstrated a protective association (ARR = 0.64; 95% CI: 0.43–0.94) indicating a 36% reduction in risk. This pattern suggests that the first three days following membrane rupture may represent a danger zone, during which the risks of ascending infection and inflammatory activation are heightened before the full benefits of medical management are realized. In contrast, the 4–7 days window likely represents a clinical success group, mothers who remained hemodynamically stable and free of overt infection long enough to complete a full course of antenatal corticosteroids and receive adequate antibiotic prophylaxis. Achieving this window may allow fetal maturation (53) while minimizing infectious complications, thereby improving neonatal outcomes. Clinically, these findings underscore the importance of intensive monitoring and stabilization during the first 72 hours after PPROM. This study may reflect context-specific factors in Tigray, such as access to prophylactic antibiotics, tocolysis, or corticosteroids, delayed presentations, or higher baseline infection rates leading to quicker progression to complications during early expectant management(25, 27, 54, 55).

A history of infection in a previous pregnancy (ARR = 1.54; 95% CI: 1.08–2.22) and pelvic pain in the current pregnancy (ARR = 2.09; 95% CI: 1.05–4.15) emerged as significant predictors of adverse neonatal outcomes. The very high proportion of adverse outcomes among women reporting foul-smelling vaginal discharge (88.9%) may further implicate underlying intrauterine infection or chorioamnionitis. This might be due to comprising the protective barrier of the fetal membranes may lead to ascending infection(56), which triggered a fetal inflammatory response(57), and increased the likelihood of respiratory distress, sepsis(9, 58), and other neonatal complications(59, 60). In resource-limited settings, delayed diagnosis and limited laboratory capacity may allow subclinical infections to progress(24), thereby worsening outcomes(3). In Tigray, where diagnostic capacity may be constrained, these two clinical features could serve as practical “red flags” during triage.

Occupational status demonstrated differential effects. Being a student was modestly associated with increased risk of adverse outcomes (ARR=1.52; 95% CI: 1.144-2.031; p=0.004) as compared to housewives. This association may reflect underlying social and demographic factors like differences in age, social stigma, limited financial autonomy, and reduced family support, which could hinder timely care-seeking and adequate antenatal follow-up. This study is consistent with broader evidence that employment status and job type proxy deeper social and behavioral factors (61–63).

Maternal nutritional and preventive factors emerged as important predictors of composite adverse perinatal outcomes. Absence of iron–folate supplementation during pregnancy increased the risk of composite adverse neonatal outcomes 63% (ARR=1.63; 95% CI: 1.153-2.29; p=0.005). This is consistent with evidence from low- and middle-income countries showing that iron deficiency anemia can worsen hypoxia-related complications and increase susceptibility to infection, impaired fetal growth(64–66), and reduce resilience to perinatal stress(67). This finding underscores the need to strengthen pre-conception and community-level nutritional strategies in post-conflict Tigray. Rather than relying solely on facility-based ANC, iron–folate distribution could be integrated into community outreach, mobile health services, and food aid programs coordinated by the Tigray Regional Health Bureau to reach vulnerable women earlier and more equitably.

Onset of labor also influenced outcomes. Induced labor was associated with a 44% reduction in adverse perinatal outcomes (ARR=0.58; 95% CI: 0 .422-0.800; p=0.001), suggesting that timely obstetric intervention following PPROM may mitigate neonatal risk(54). Induction may lower the composite of adverse perinatal outcomes by shortening exposure to ascending infection once a reasonable gestational age and steroid course have been achieved, and this is supported by management guideline recommendations (54, 68).

Collectively, these findings suggest that PPROM contributes to adverse neonatal outcomes through intertwined pathways of prematurity and infection, both of which are exacerbated in settings with constrained neonatal intensive care capacity. Delayed care-seeking after membrane rupture was associated with a notably higher proportion of composite adverse neonatal outcomes (40.2% among those presenting after >6 hours). In the context of Tigray, this delay is unlikely to be merely individual-level behavior but may reflect broader structural barriers created by the recent armed conflict. The destruction of transportation networks, disruption of referral systems, shortages of healthcare personnel, fuel scarcity, and reduced functionality of peripheral health facilities likely impeded timely access to emergency obstetric care.

## Conclusion

This prospective cohort study demonstrates that preterm premature rupture of membrane (PPROM) is the most powerful independent predictor of composite adverse perinatal outcome in public hospitals of Tigray Region. Neonates born to mothers with PPROM were more than seven times more likely to experience at least one serious adverse outcome compared to those without PPROM. The burden was particularly pronounced among preterm and small for gestational age neonates and those requiring immediate resuscitation or NICU admission.

The first 72 hours following membrane rupture emerged as a critical high-risk period, during which the likelihood of adverse outcomes significantly increased. Previous pregnancy related infection, absence of iron folate supplementation, pelvic pain during pregnancy, and prolonged latency (1–3 days) further amplified the risk, whereas induced labor showed a protective effect.

The findings highlight PPROM as both clinical emergency and a health system challenge in post conflict, resource limited settings. Strengthening infection screening and prevention, ensuring iron folate supplementation, improving early detection of warning symptom, and implementing structural monitoring protocols during the early latency period could substantially reduce preventable neonatal morbidity and mortality. Targeted, context specific interventions focused on early stabilization and timely obstetric decision making are essential to improve neonatal survival.

## Limitation of study

This study is observational in nature and the findings demonstrate association rather than direct causal relationship. Although multivariable adjustment and robust standard errors were applied to address confounding and under-dispersion, residual confounding can’t be excluded. Additionally, unmeasured clinical variables and contextual factors may have influenced the observed relationship.

**Table 1:**
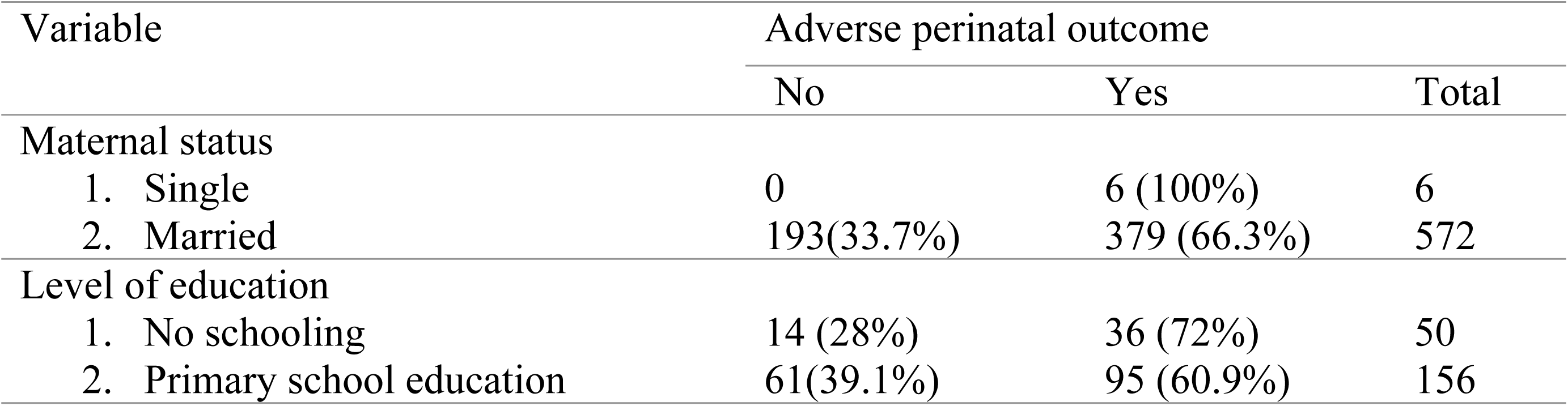
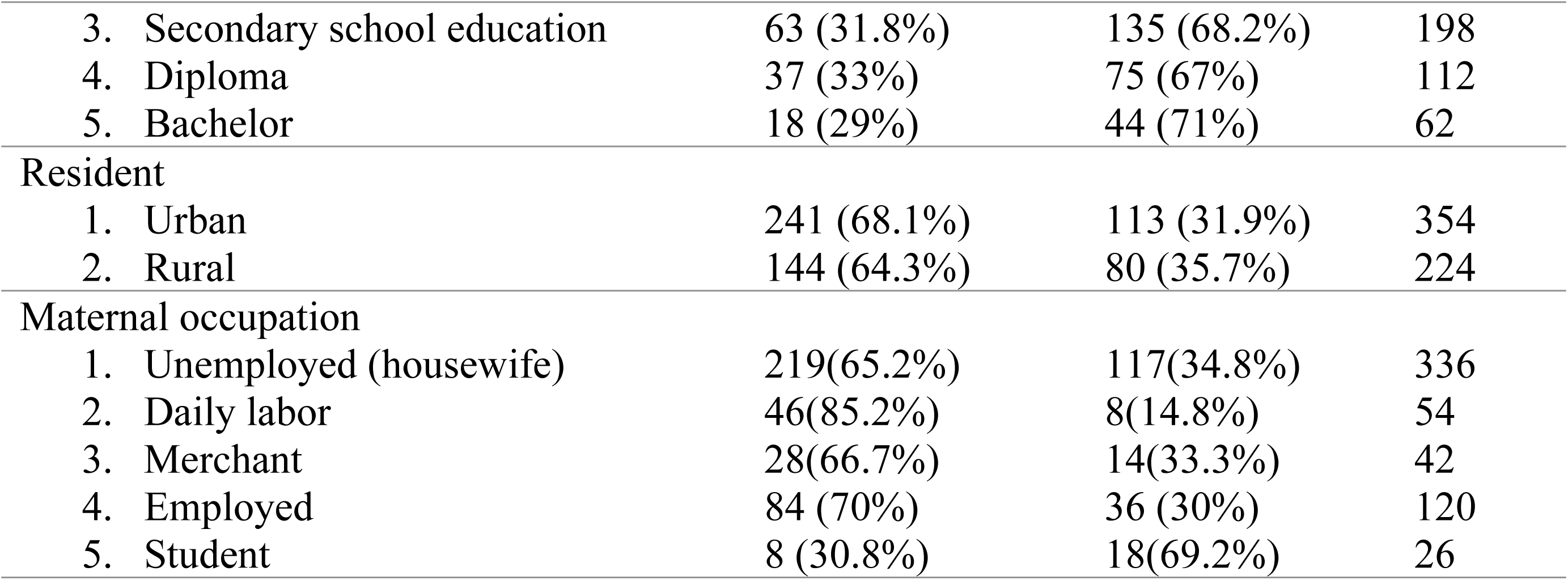
Socio-demographic characteristics of mothers and composite adverse perinatal outcome among study participants in Tigray region, northern Ethiopia.

| Variable | Adverse perinatal outcome |  |  |
| --- | --- | --- | --- |
|  | No | Yes | Total |
| Maternal status |  |  |  |
| 1. Single | 0 | 6 (100%) | 6 |
| 2. Married | 193(33.7%) | 379 (66.3%) | 572 |
| Level of education |  |  |  |
| 1. No schooling | 14 (28%) | 36 (72%) | 50 |
| 2. Primary school education | 61(39.1%) | 95 (60.9%) | 156 |
| 3. Secondary school education | 63 (31.8%) | 135 (68.2%) | 198 |
| 4. Diploma | 37 (33%) | 75 (67%) | 112 |
| 5. Bachelor | 18 (29%) | 44 (71%) | 62 |
| Resident |  |  |  |
| 1. Urban | 241 (68.1%) | 113 (31.9%) | 354 |
| 2. Rural | 144 (64.3%) | 80 (35.7%) | 224 |
| Maternal occupation |  |  |  |
| 1. Unemployed (housewife) | 219(65.2%) | 117(34.8%) | 336 |
| 2. Daily labor | 46(85.2%) | 8(14.8%) | 54 |
| 3. Merchant | 28(66.7%) | 14(33.3%) | 42 |
| 4. Employed | 84 (70%) | 36 (30%) | 120 |
| 5. Student | 8 (30.8%) | 18(69.2%) | 26 |

**Table 2:** Previous obstetric history of mothers and composite adverse perinatal outcome among study participants in Tigray region, northern Ethiopia.

| Variable | Adverse perinatal outcome |  |  |
| --- | --- | --- | --- |
|  | No | Yes | Total |
| History of previous preterm birth |  |  |  |
| 1. Yes | 20 (58.8%) | 34 (41.2%) | 54 |
| 2. No | 365(67.1%) | 179 (32.9%) | 544 |
| History of previous PPRM |  |  |  |
| 1. Yes | 26 (72.2%) | 10 (27.8%) | 36 |
| 2. No | 359 (66.2%) | 183 (33.8%) | 542 |
| History of previous still birth |  |  |  |
| 1. Yes | 179 (32.7%) | 369 (67.3%) | 548 |
| 2. No | 14 (46.7%) | 16 (53.3%) | 30 |
| History of previous miscarriage/abortion |  |  |  |
| 1. Yes | 52 (61.9%) | 32 (38.1%) | 84 |
| 2. No | 333 (67.4%) | 161 (32.6%) | 494 |
| History of previous pregnancy complication |  |  |  |
| 1. Yes | 22 (52.4%) | 20 (47.6%) | 42 |
| 2. No | 363 (67.7%) | 173 (32.3%) | 536 |
| Type of complication |  |  |  |
| 1. High blood pressure | 4 (66.7%) | 2 (33.3%) | 6 |
| 2. Gestational diabetics | 0 | 2 (100%) | 2 |
| 3. Excessive bleeding | 8(57.1%) | 6 (42.9%) | 14 |
| 4. Fetal distress | 8 (66.7%) | 4 (33.3%) | 12 |
| 5. Long labor | 0 | 2 (100) | 2 |
| 6. Other1 | 10 (50%) | 10 (50%) | 20 |
| History of previous CS delivery |  |  |  |
| 1. Yes | 28 (58.3%) | 20 (41.7%) | 48 |
| 2. No | 357 (67.4%) | 173 (32.6%) | 530 |
| Type of C/S |  |  |  |
| 1. Planned | 2 (50%) | 2 (50%) | 4 |
| 2. Emergency or emergency | 26 (59.1%) | 18 (40.9%) | 44 |
| Previous pregnancy affected by infection |  |  |  |
| 1. Yes | 2 (20%) | 8 (80%) | 10 |
| 2. No | 383 (67.4%) | 185 (32.6%) | 568 |
| Regular ANC in previous pregnancy |  |  |  |
| 1. Yes | 173 (69.2%) | 77 (30.8%) | 250 |
| 2. No | 212 (64.6%) | 116 (35.4%) | 328 |
| Medical treatment previous pregnancy |  |  |  |
| 1. Yes antibiotic | 33 (50%) | 33 (50%) | 66 |
| 2. Yes steroid | 10 (38.5%) | 16 (61.5%) | 26 |
| 3. Yes, other treatment | 6 (75%) | 2 (25%) | 8 |
| 4. No | 348 (69%) | 156 (32%) | 540 |
| Bleeding during previous pregnancy |  |  |  |
| 1. Yes | 16 (61.5%) | 10 (38.5%) | 26 |
| 2. No | 369 (66.8%) | 183 (33.2%) | 552 |
| Previous NICU admission |  |  |  |
| 1. Yes | 16 (66.7%) | 8 (33.3%) | 24 |
| 2. No | 369 (66.6%) | 185 (33.4%) | 554 |
| Reason for NICU admission |  |  |  |
| 1. Low birth weight | 8 (66.7%) | 4 (33.3%) | 12 |
| 2. Unable to suck | 8 (80%) | 2 (20%) | 10 |
| 3. Preterm birth | 6 (60%) | 4 (40%) | 10 |
| 4. Hypothermia | 2 (33.3%) | 4 (66.7%) | 6 |
| 5. Meconium aspiration syndrome | 4 (100%) | 0 | 4 |
| 6. Neonatal sepsis | 2 (100%) | 0 | 2 |
| Previous low birth weight (below 2.5 kg) |  |  |  |
| 1. Yes | 17 (60.7%) | 11 (39.3%) | 28 |
| 2. No | 368 (66.9%) | 182 (33.1%) | 550 |
| Baby born with any congenital conditions |  |  |  |
| 1. Yes | 4 | 0 | 4 |
| 2. No | 381 (66.4%) | 193 (33.6%) | 574 |
| Medical conditions before becoming pregnant |  |  |  |
| 1. Yes | 6 (100%) | 0 | 6 |
| 2. No | 379 (66.3%) | 193 (33.7%) | 572 |
| Pre-pregnancy supplements |  |  |  |
| 1. Yes | 169 (71.6%) | 67 (28.4%) | 236 |
| 2. No | 216 (63.2%) | 126 (36.8%) | 342 |
| Complications with placenta position or function in previous pregnancies |  |  |  |
| 1. Yes | 6 (50%) | 6 (50%) | 12 |
| 2. No | 379 (67%) | 187 (33%) | 566 |

Other1: Abortion, CPD, Incomplete placenta

**Table 3:** Obstetrics and clinical characteristics of Current pregnancy on composite adverse perinatal outcome among study participants in Tigray region, northern Ethiopia.

| Variable | Adverse perinatal outcome |  |  |
| --- | --- | --- | --- |
|  | No | Yes | Total |
| MUAC of mothers |  |  |  |
| 1. $\geq 23$ cm | 281(67.2%) | 137(32.8) | 418 |
| 2. <23 cm | 104(65%) | 56(35%) | 160 |
| Experience of fluid leakage before labor |  |  |  |
| 1. Yes | 330 (64%) | 186 (36%) | 516 |
| 2. No | 55(88.7%) | 7 (11.3%) | 62 |
| Current diagnosis with PPRM |  |  |  |
| 1. Yes | 117 (40.6%) | 171 (59.4%) | 288 |
| 2. No | 268(92.4%) | 22 (7.6%) | 290 |
| Hospitalized immediately after PPRM |  |  |  |
| 1. Yes | 111(39.9%) | 167 (60.1%) | 278 |
| 2. No | 6 (75%) | 2 (25%) | 8 |
| Gestational age at rupture of membrane |  |  |  |
| 1. 28-33<sup>+6</sup> weeks | 19 (22.1%) | 67 (77.9%) | 86 |
| 2. 34-36<sup>+6</sup> weeks | 96 (47.5%) | 106 (52.5%) | 202 |
| 3. 37 weeks and above | 270 (93.1%) | 20 (6.9%) | 290 |
| Time from rupture of membrane to seeking care |  |  |  |
| 1. With in 1 hour | 97 (71.3%) | 39(28.7%) | 136 |
| 2. 1-3 hours | 111(67.7%) | 53 (32.3%) | 164 |
| 3. 4-6 hours | 79 (69.3 %) | 35 (30.7%) | 114 |
| 4. More than 6 hours | 98 (59.8%) | 66 (40.2%) | 164 |
| Received treatment after PPRM |  |  |  |
| 1. Yes | 115 (40.8%) | 167 (59.2%) | 282 |
| 2. No | 2(33.3%) | 4(66.4%)) | 6 |
| Type of treatment from card |  |  |  |
| 1. Antibiotics | 115 (41.1%) | 165 (58.9%) | 280 |
| 2. Corticosteroids | 100 (41.3%) | 142 (58.7%) | 242 |
| 3. Tocolytics | 15 (20.3%) | 59 (79.7%) | 74 |
| 4. Other | 2 (16.7%) | 10 (83.3/%) | 12 |
| Satisfaction by health care givers |  |  |  |
| 1. Good | 93 (64.6%) | 51(35.4%) | 144 |
| 2. Average | 4 (50%) | 4(50%) | 8 |
| 3. Excellent | 288(67.6%) | 138 (32.4%) | 426 |
| Birth interval |  |  |  |
| 1. >=2 years | 232(71.6%) | 92(28.4%) | 324 |
| 2. <2 years | 153(60.2) | 101(39.8%) | 254 |
| Parity |  |  |  |
| 1. Primiparous | 148(59.7%) | 100(40.3%) | 248 |
| 2. Multiparous | 237(71.8%) | 93 (28.2%) | 330 |
| Antenatal visits in the current pregnancy |  |  |  |
| 1. Yes | 383 (66.7%) | 191 (33.3%) | 574 |
| 2. No | 2 (50%) | 2 (50%) | 4 |
| Place of ANC attendance |  |  |  |
| 1. Health center | 281 (67.5%) | 135 (32.5%) | 416 |
| 2. General hospital | 233(65.8%) | 121 (34.2%) | 354 |
| 3. Primary hospital | 82 (63.1%) | 48(36.9%) | 130 |
| 4. Specialized hospital | 11(36.7%) | 19(63.3%) | 30 |
| Influences your decision to seek ANC |  |  |  |
| 1. Advice from family members | 286(66.2%) | 146(33.8%) | 432 |
| 2. Personal health concerns | 265(62.2%) | 161(37.8%) | 4 |
| 3. Doctors' recommendation | 189 (69.5%) | 83(30.5%) | 432 |
| 4. Financial ability | 96 (70.6%) | 40 (29.4%) | 428 |
| 5. Transportation access | 55(72.4%) | 21 (27.6%) | 76 |
| 6. Other | 8(100%) | 0 | 8 |
| Accessibility |  |  |  |
| 1. Very easy | 197 (63.1%) | 115 (36.9%) | 312 |
| 2. Somehow easy | 140 (68.6%) | 64 (34.4%) | 204 |
| 3. Difficult | 42 (77.8%) | 12 (22.2%) | 54 |
| 4. Very difficult | 6 (75%) | 2 (25%) | 8 |
| Primary means of transportation |  |  |  |
| 1. Walking | 132(70.2%) | 56 (29.8%) | 188 |
| 2. Personal vehicle | 10 (50%) | 10 (50%) | 20 |
| 3. Public transport | 237(66.2%) | 121(33.8%) | 358 |
| 4. Other | 6(50%) | 6(50%) | 12 |
| Current Iron folate supplementation |  |  |  |
| 1. Yes | 360(67.9%) | 170(32.1%) | 530 |
| 2. No | 25(54.3%) | 21(45.7%) | 46 |
| TT Vaccination in current pregnancy |  |  |  |
| 1. Yes | 301(64.6%) | 165(35.4%) | 466 |
| 2. No | 82(74.5%) | 28(25.5%) | 110 |
| Experience of UTI in this pregnancy |  |  |  |
| 1. Yes | 74(77.1%) | 22(22.9%) | 99 |
| 2. No | 311(64.5%) | 171(35.5%) | 482 |
| Experience of recent accident /trauma/ |  |  |  |
| 1. Yes | 12(85.7%) | 2(14.3%) | 14 |
| 2. No | 373(66.1%) | 191(33.9%) | 564 |
| Fetal presentation |  |  |  |
| 1. Cephalic | 346(68.7%) | 158(31.3%) | 504 |
| 2. Non-cephalic | 39(52.7%) | 35(47.3%) | 74 |
| APH in current pregnancy |  |  |  |
| 1. Yes | 13(81.3%) | 3(18.8%) | 16 |
| 2. No | 373(66.2%) | 190(33.8%) | 563 |
| Current Diagnosis with pre-eclampsia/eclampsia |  |  |  |
| 1. Yes | 6(37.5%) | 10(62.5%) | 16 |
| 2. No | 379(67.4%) | 183(32.6%) | 562 |
| Current HGB level |  |  |  |
| 1. <11g/dl | 22(68.8%) | 10(31.3%) | 32 |
| 2. ≥11g/dl | 363(66.5%) | 183(33.5%) | 546 |
| Vaginal bleeding in current pregnancy |  |  |  |
| 1. Yes | 5(50%) | 5(50%) | 10 |
| 2. No | 380(66.9%) | 188(33.1%) | 568 |
| Chronic cough in this pregnancy |  |  |  |
| 1. Yes | 26(76.5%) | 8(23.5%) | 34 |
| 2. No | 359(66%) | 185(34%) | 544 |
| Gestational hypertension in this pregnancy |  |  |  |
| 1. Yes | 12(54.5%) | 10(45.5%) | 22 |
| 2. No | 373(67.1%) | 182(32.9%) | 556 |
| Carrying heavy objects in this pregnancy |  |  |  |
| 1. Yes | 60(76.9%) | 18(23.1%) | 78 |
| 2. No | 325(65%) | 175(35%) | 500 |

**Table 4:** Maternal health-seeking behavior and composite adverse perinatal outcome among study participants in Tigray region, northern Ethiopia.

| Variable | Adverse perinatal outcome |  |  |
| --- | --- | --- | --- |
|  | No | Yes | Total |
| Feel comfortable discussing health |  |  |  |
| 1. Yes, comfortable | 383(67%) | 189(33%) | 572 |
| 2. No, uncomfortable | 2(33.3%) | 4(66.7%) | 6 |
| Skipped ANC appointment |  |  |  |
| 1. Yes | 145(64.2%) | 81(35.8%) | 226 |
| 2. No | 240(68.2%) | 112(31.8%) | 352 |
| Reason of ANC skipping |  |  |  |
| 1. Financial issue | 107(63.7%) | 61(36.3%) | 168 |
| 2. Lack of transportation | 125(66.5%) | 63(33.5%) | 188 |
| 3. Distance to health care facility | 86(64.2%) | 48(35.8%) | 134 |
| 4. Fear of diagnosis | 18(42.9%) | 24(57.1%) | 42 |
| 5. Believe in alternative treatments | 4(50%) | 4(50%) | 8 |
| 6. Family opposition | 8(50%) | 8(50%) | 16 |
| Family support for seeking health care |  |  |  |
| 1. Yes | 379(66.5%) | 191(33.5%) | 570 |
| 2. No | 6 (75%) | 2 (25%) | 8 |
| Satisfaction with care information |  |  |  |
| 1. Yes | 383(67%) | 189(33%) | 572 |
| 2. No | 2(33.3%) | 4(66.7%) | 6 |
| You reason to choose this healthcare service |  |  |  |
| 1. Proximity to home | 175(69.4%) | 77(30.6%) | 252 |
| 2. Quality of care | 301(66.6%) | 151(33.4%) | 452 |
| 3. Reputation of healthcare provider | 61(62.2%) | 37(37.8%) | 98 |
| 4. Cost | 89(64.5%) | 49(35.5%) | 138 |
| 5. Recommendation from family/friend | 66(55%) | 54(45%) | 120 |
| 6. Other | 32(59.3%) | 22(40.7%) | 55 |

Other: *Personal choice*

**Table 5:** Infection-related clinical factors by composite adverse perinatal outcome among study participants in Tigray region, northern Ethiopia.

| Variable | Adverse perinatal outcome |  |  |
| --- | --- | --- | --- |
|  | No | Yes | Total |
| PV over examination ($\geq 5$times) | | | |
| 1. Yes | 106(65.4%) | 56(34.6%) | 162 |
| 2. No | 279(67.1%) | 137(32.9%) | 416 |
| Foul smelling of vaginal discharge |  |  |  |
| 1. Yes | 2(11.1%) | 16(88.9%) | 18 |
| 2. No | 383(68.4%) | 177(31.6%) | 560 |
| Experience of pelvic pain this pregnancy |  |  |  |
| 1. Yes | 69(50%) | 6 (50%) | 12 |
| 2. No | 379(67%) | 187(33%) | 466 |
| Clarity explanation about treatment options |  |  |  |
| 1. Yes | 305(64.3%) | 169(35.7%) | 474 |
| 2. No | 80(76.9%) | 24(23.1%) | 104 |
| Explanation on potential risk to your baby |  |  |  |
| 1. Yes | 290(66.2%) | 148(33.8%) | 438 |
| 2. No | 95(67.9%) | 45(32.1%) | 140 |
| Provide contact information on emergency |  |  |  |
| 1. Yes | 325(65.8%) | 169(34.2%) | 494 |
| 2. No | 60(71.4%) | 24(28.6%) | 84 |
| Baby receive antibiotics post-birth |  |  |  |
| 1. Yes | 5(5.3%) | 89(94.7%) | 94 |
| 2. No | 380(79.8%) | 96(20.2%) | 476 |

**Table 6:** Delivery characteristics and immediate neonatal outcomes by composite adverse perinatal outcome status among study participants in Tigray region, northern Ethiopia.

| Variable | Adverse perinatal outcome |  |  |
| --- | --- | --- | --- |
|  | No | Yes | Total |
| Latency period (between ROM & delivery) |  |  |  |
| 1. Less than 24 hours | 173(77.9%) | 49(22.1%) | 222 |
| 2. 1-3 days | 120(63.8%) | 68(36.2%) | 188 |
| 3. 4-7 days | 36(64.3%) | 20(35.7%) | 56 |
| 4. More than 7 days | 56(50%) | 56(50%) | 112 |
| Complication during latency period |  |  |  |
| 1. Yes | 353(68.7%) | 161(31.3%) | 514 |
| 2. No | 32(50%) | 32(50%) | 64 |
| Type of complications |  |  |  |
| 1. Infection (e.g. chorioamnionitis) | 0 | 8(100%) | 8 |
| 2. Bleeding | 0 | 2(100%) | 2 |
| 3. Increased uterine contraction | 2(33.3%) | 4(66.7%) | 6 |
| 4. Placental problems | 0 | 2(100%) | 2 |
| 5. C/S delivery | 28(70%) | 12(30%) | 40 |
| 6. Abnormal fetal presentation | 14(77.8%) | 4(22.2%) | 16 |
| 7. Other1 | 4(25%) | 12(75%) | 16 |
| Undergo lab test |  |  |  |
| 1. Yes | 377(66.6%) | 189(33.4%) | 566 |
| 2. No | 8(66.7%) | 4(33.3%) | 12 |
| Onset of labor |  |  |  |
| 1. Spontaneous | 249(61.3%) | 157(38.7%) | 406 |
| 2. Induced | 136(79.1%) | 36 (20.9%) | 172 |
| Sex of neonate |  |  |  |
| 1. Male | 220(66.3%) | 112(33.7%) | 332 |
| 2. Female | 165(67.1%) | 81(32.9%) | 246 |
| Mode of delivery |  |  |  |
| 1. Vaginal | 302(66.8%) | 150(33.2%) | 452 |
| 2. C/S | 57(62%) | 35(38%) | 92 |
| 3. Vaginal with instrument | 26(76.5%) | 8(23.5%) | 34 |
| Who delivered you |  |  |  |
| 1. Midwifery | 323(67%) | 159(33%) | 482 |
| 2. Nurse | 1(50%) | 1(50%) | 2 |
| 3. Gyn/obstetrician | 40(57.1%) | 30(42.9%) | 70 |
| 4. Emergency surgeon | 51(67.1%) | 25(32.9%) | 76 |
| Gestational age at delivery in weeks |  |  |  |
| 1. 28-33+6 weeks | 6(15.8%) | 32(84.2%) | 38 |
| 2. 34-36+6 weeks | 41(24.7%) | 125(75.3%) | 166 |
| 3. 37 weeks and above | 338(90.4%) | 36(9.6%) | 374 |
| Gestational age with birth weight |  |  |  |
| 1. Small for gestational age | 26(31.7%) | 56(68.3%) | 82 |
| 2. Appropriate for gestational age | 307(70.4%) | 129(29.6%) | 436 |
| 3. Large for gestational age | 52(86.7%) | 8(13.3%) | 60 |
| Resuscitation intervention during delivery |  |  |  |
| 1. Yes | 21(19.4%) | 87(80.6%) | 108 |
| 2. No | 358(82.1%) | 8(17.9%) | 436 |
| Complication during delivery |  |  |  |
| 1. Yes | 13(29.5%) | 31(70.5%) | 44 |
| 2. No | 372(69.7%) | 162(30.3%) | 534 |
| Type of complication |  |  |  |
| 1. Placental abruptio/previa | 10(38.5%) | 16(61.5%) | 26 |
| 2. Meconium aspiration syndrome | 4(20%) | 16(80%) | 20 |

**Table 7:** Perinatal outcome by composite adverse perinatal outcome status among study participants in Tigray region, northern Ethiopia.

| Variable | Adverse perinatal outcome |  |  |
| --- | --- | --- | --- |
|  | No | Yes | Total |
| APGAR score 1<sup>st</sup> minute |  |  |  |
| 1. 4-7 | 18(18.4%) | 80(81.6%) | 98 |
| 2. 7-10 | 367(77.8%) | 105(22.2%) | 472 |
| APGAR score 5<sup>th</sup> minute |  |  |  |
| 1. 4-7 | 2(4%) | 48(96%) | 50 |
| 2. 7-10 | 383(73.7%) | 137(26.3%) | 520 |
| Baby admitted to NICU |  |  |  |
| 1. Yes | 12(9.8%) | 110(90.2%) | 122 |
| 2. No | 373(83.3%) | 75(16.7%) | 448 |
| Reason for admission |  |  |  |
| 1. Respiratory distress syndrome | 5(8.1%) | 57(91.9%) | 62 |
| 2. Hypothermia | 2(20%) | 8(80%) | 10 |
| 3. Neonatal sepsis | 0 | 2(100%) | 2 |
| 4. Low birth weight | 8(9.8%) | 74(90.2%) | 82 |
| 5. Jaundice | 0 | 4(100%) | 4 |
| 6. Feeding difficulties | 3(13.6%) | 19(86.4%) | 22 |
| 7. Meconium aspiration syndrome | 2(10%) | 18(90%) | 20 |
| 8. Other5 | 1(6.3%) | 15(92.7%) | 16 |
| Baby respiratory support during delivery |  |  |  |
| 1. Yes | 8(9.1%) | 80(90.9%) | 88 |
| 2. No | 4(14.3%) | 24(85.3%) | 28 |
| Baby receive supplemental oxygen |  |  |  |
| 1. Yes | 12(12%) | 88(88%) | 100 |
| 2. No | 373(79.4%) | 97(20.6%) | 470 |

**Table 8:** Crude and Adjusted results of Modified Poisson Regression Analysis for the effect of preterm premature rupture of membranes on composite adverse perinatal outcome and its predictors

| Characteristics | Variable Category | CRR | ARR (95% CI) | p-value |
| --- | --- | --- | --- | --- |
| Maternal age in years |  | 0.979 | 0.98(0.960-1.00) | 0.144 |
| Maternal occupation | Unemployed (housewife) | Ref | Ref. | 1 |
|  | Daily labor | 0.425 | 0.47(0.217-1.03) | 0.059 |
|  | Merchant | 0.957 | 1.38(0.837-2.28) | 0.206 |
|  | Employed | 0.862 | 0.96(0.726-1.266) | 0.766 |
|  | Student | 1.988 | **1.52(1.144-2.031)** | **0.004\*\*** |
| History of previous still birth | Yes | 1.429 | 1.55(0.936-2.563) | 0.089 |
|  | No | Ref. | 1 |  |
| History of previous pregnancy complications | Yes | 1.475 | 1.19(0.369-2.952) | 0.937 |
|  | No | Ref. | 1 |  |
| History of previous CS delivery | Yes | 1.276 | 1.32(0. .912-2.012) | 0.132 |
|  | No | Ref. | 1 |  |
| History of previous pregnancy infection | Yes | 2.456 | **1.54(1.075-2.215)** | **0.019\*\*** |
|  | No | Ref. | 1 |  |
| Previous Pregnancy placental complications | Yes | 1.513 | 1.19(0.429-3.3016) | 0.738 |
|  | No | Ref. | 1 |  |
| Pre-pregnancy supplementation | Yes | Ref. | 1 |  |
|  | No | 1.298 | 1.22(0.99-1.1.52) | 0.063 |
| Current pregnancy diagnosis with PPRM | Yes | 7.827 | **7.22(4.73-11.03)** | **0.001\*\*** |
|  | No | Ref. | 1 |  |
| Parity | Multiparous | Ref. | 1 |  |
|  | Primiparous | 1.430 | 1.08(0.827-1.35) | 0.659 |
| Current pregnancy Iron folate supplementation | Yes | Ref. | 1 |  |
|  | No | 1.494 | **1.63(1.153-2.29)** | **0.005\*\*** |
| Experience UTI in current pregnancy | Yes | 0.646 | 0.80(0.545-1.195) | 0.285 |
|  | No | Ref. | 1 |  |
| Fetal Presentation | Cephalic | Ref. | 1 |  |
|  | Non-cephalic | 1.509 | 1.22(0.919-1.632) | 0.172 |
| Current Diagnosis with pre-eclampsia /eclampsia | Yes | 1.919 | 1.20(0.796-1.836) | 0.374 |
|  | No | Ref. | 1 |  |
| Vaginal bleeding in the current pregnancy | Yes | 1.510 | 0.48(0.222-1.036) | 0.061 |
|  | No | Ref. | 1 |  |
| Carrying heavy object in the current pregnancy | Yes | 0.659 | 0.98(0.631-1.521) | 0.928 |
|  | No | Ref. | 1 |  |
| Experiencing of pelvic pain in current pregnancy | Yes | 1.513 | **2.09(1.053-4.152)** | **0.035\*\*** |
|  | No | Ref. | 1 |  |
| Healthcare providers explained well (clarity explanation) | Yes | Ref. | 1 |  |
|  | No | 0.647 | 0.83(0.809-1.124) | 0.225 |
| Latency period in hours (time between ROM and delivery) | Less than 24 hours | Ref. | Ref. |  |
|  | 1-3 days | 1.64 | **1.41 (1.099-1.812)** | **0.007\*\*** |
|  | 4-7 days | 1.62 | **0.64(0.433-0.943)** | **0.024\*\*** |
|  | More than 7days | 2.26 | 0.99(.732-1.35) | 0.97 |
| Experience of any complications in the latency period | Yes | 1.596 | 1.33(0.994-1.792) | 0.055 |
|  | No | Ref. | 1 |  |
| Onset of labor | Spontaneous | Ref. | 1 |  |
|  | Induced | 0.541 | **0.58(0.422-0.800)** | **0.001\*\*** |

## Abbreviations

ARR: Adjusted Relative Ratio
CRR: Crude Relative Ratio
CI: Confidence Interval
NICU: Neonatal Intensive Care Unit
PPROM: preterm premature rupture of membrane

## Declarations

### Ethical approval and consent to participate

Ethical approval was obtained from the Health Research Ethical Review Committee (HRERC) of the Aksum University College of Health Sciences and comprehensive Specialized hospital (CHS-SRH), Ethiopia (approval number 075/2024) on 23 November2024. Approval was secured before the initiation of data collection. After that an official support letter was secured from Tigray Regional Health Bureau. Written informed consent was obtained from all participants prior to their inclusion in the study. Each participant received detailed information regarding the purpose of the research, study procedure, potential risks and benefits, and their right to withdraw at any stage without any impact on the care they received. Participant confidentiality was strictly maintained through the use of unique identification codes in place of personal identifier. All data were stored securely and were accessible only to authorized members of the research teams. This investigation was conducted as an observational prospective cohort study and did not involve any interventional procedure and therefore, clinical trial registration was not required.

### Consent for publication

Not applicable

### Availability of data and materials

All relevant data are within the manuscript and its Supporting Information files.

### Authors’ contributions

As the study’s lead investigator, GTW Conceptualization, Data curation, Formal analysis, Investigation, Methodology, Project administration, Resources, Software, Supervision, Validation Visualization, Writing – original draft, Writing – review & editing. AA, GN, BA, HM, FA, HG, TG, NA, TGH, TTA Data curation, Formal analysis, Investigation, Methodology, Software, Supervision, Validation, Visualization

